# Polydrug use and recreational motives are associated with lower posttraumatic symptoms in survivors of a mass trauma event

**DOI:** 10.64898/2026.10.29.26366805

**Authors:** Noa Magal, Ophir Netzer, David H. Epstein, Yavin Shaham, Dimitris Repantis, Shaul Lev-Ran, Amir Harduf, Roee Admon, Roy Salomon

## Abstract

**Objective:** Drug use after trauma is common, but polydrug use patterns, motives, and associations with subsequent symptoms are poorly understood. We examined these associations among mass-casualty terror attack survivors, 72% of whom used drugs immediately before trauma exposure.

**Methods:** Survivors of the October 7, 2023 Nova Festival attack reported use of six drug types (alcohol, cannabis, ketamine, stimulants, MDMA, psychedelics) and motives (avoidance, emotional regulation, self-medication, sleep, recreation) longitudinally at three timepoints (TP): 1-6 months (N=772), 8-12 months (N=453), and 17-24 months (N=302) post-attack. Separate latent class analyses of TP2 data characterized drug use, motives, and their combination. Linear regression and mixed-effects models examined associations between class membership and PTSD Checklist for DSM-5 (PCL-5) scores, adjusting for age, sex, time since trauma, and TP1 PCL-5 scores.

**Results:** At TP2, 70% reported polydrug use, 17% used one drug, and 13% reported no use. The high drug-load class (mean 4.6 drug types) had lower PCL-5 scores than the low drug-load class (mean 1.5 drug types); this difference was evident among participants using ≥4 drug types and persisted through TP3. The self-medication-motive class had higher PCL-5 scores than the recreational-motive class. In joint analyses, recreational-motive users at both low and high drug loads had lower PCL-5 scores than participants reporting self-medication motives and primarily cannabis use.

**Conclusions:** Polydrug use was common among the mass-trauma survivors in our study. Self-medication motives were associated with higher PTSD symptom severity than recreational motives. Unexpectedly, higher polydrug use was associated with lower PTSD symptom scores.

## Introduction

Posttraumatic stress disorder (PTSD) is associated with the use of potentially addictive drugs; causes of the association may include self-medication for PTSD symptoms (1, 2). Epidemiological studies suggest that such self-medication is often ineffective (3), and if drug use transitions into a substance use disorder (SUD), the prognosis for comorbid SUD/PTSD is poorer than for either alone (4).

Research on PTSD and SUDs has usually focused on drugs other than the psychedelics and entactogens that have moved toward clinical use for treatment or prevention of PTSD (5–7). Controlled trials of MDMA-assisted therapy and ketamine have reported promising effects on posttraumatic symptoms (6, 8), generating clinical, regulatory, and public interest in psychedelic medicine (5, 9). However, outside supervised treatment protocols, trauma survivors already use these drugs on their own, at uncontrolled doses, with other drugs, and in settings unlike those of clinical trials. These naturalistic patterns of drug use have rarely been examined together with motives for use in the context of emergent PTSD symptoms after trauma exposure.

A rare opportunity to address this arose in the aftermath of the October 7, 2023 attack at the Nova music festival in Israel. Several thousand festival attendees were directly exposed to a mass-casualty terror attack on a single day, creating a large and identifiable survivor population that we have followed prospectively with repeated assessments of drug use (though not SUD symptoms), motives for use, and posttraumatic symptoms for two years since the event (10–12).

We previously found in this sample that attendees who had taken MDMA shortly before the attack, thereby experiencing the attack under the influence of MDMA, had more favorable posttraumatic outcomes than those who had not used any drugs that day. Their outcomes over the first four months after the attack included lower psychological distress, greater social interaction, and better sleep, accompanied by a perception that the drug use had been helpful during the event (11). In contrast, cannabis and/or alcohol use shortly before the attack was associated with greater PTSD symptoms, psychological distress and poorer sleep, compared to use of no drugs (11).

In the current report we characterize motives for, and patterns of, ongoing drug use among Nova festival survivors assessed longitudinally 8 to 24 months after the attack. Using latent class analysis (LCA), we identify classes based on (a) types of drugs used, (b) motives for drug use, and (c) their combination. We then examine class differences in severity of emergent PTSD symptoms. Based on prior work(e.g. 3), we expected self-medication motives to be associated with higher PTSD symptom severity.

## Methods

### Participants and data collection

Participants were recruited from a longitudinal study of survivors directly exposed to the October 7, 2023 attack at the Nova music festival in Israel (10). From an estimated population of 3,710 survivors (66% male; 76% aged 18–24 years), 1,268 were enrolled (772 men, 495 women, 1 unspecified; mean age=27.17 years, SD=6.75). Recruitment was conducted collaboratively with Safeheart (a nonprofit organization supporting Nova survivors), support groups on social media, and word of mouth. Data were collected between November 2, 2023 and September 26, 2025. All participants provided informed consent. The study was approved by the Institutional Ethics Committee of the University of Haifa (Approval No. 374/23). Assessments were performed longitudinally at three post-event timepoints (TPs): TP1 (1-6 months), TP2 (8-12 months), and TP3 (17-24 months) (**Figure 1A**). The main analyses here included participants aged ≥18 years who completed both TP1 and TP2 assessments. Longitudinal analyses were restricted to participants with data available at TP3. **Table 1** shows demographic and clinical characteristics of the sample.

**Figure 1.**
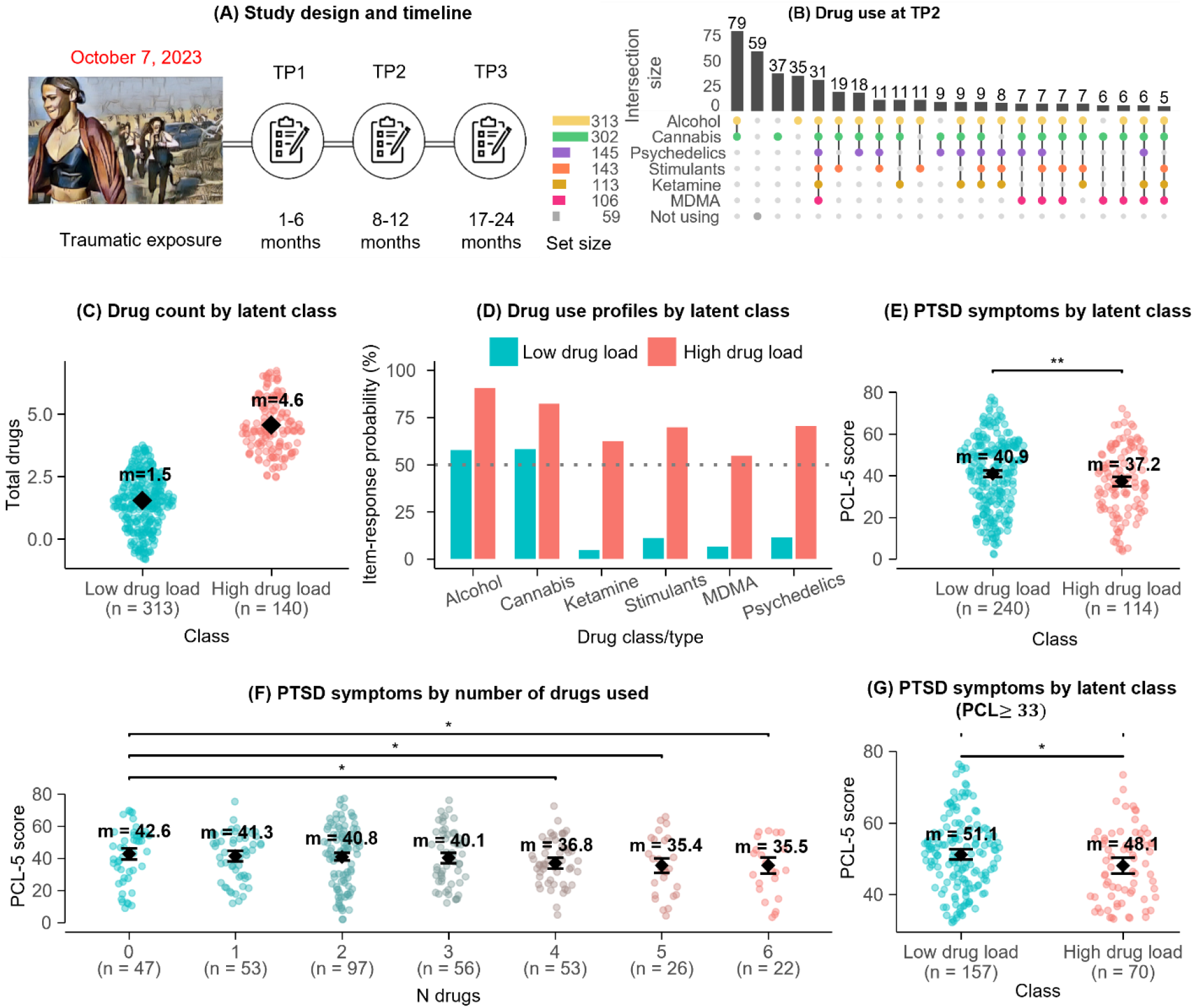
Study design, drug use patterns, and PTSD symptoms at TP2. **(A) Study design and timeline.** Assessments relative to the October 7, 2023 traumatic exposure: TP1 (1–6 months post-event), TP2 (8–12 months) and TP3 (17–24 months). **(B) Drug use at TP2.** UpSet plot of the drug combinations used in the month before TP2 (n=453). Vertical bars show the number of participants with each combination (combinations with at least 5 participants are shown); the dot matrix shows which drugs form each combination; horizontal bars on the left show the number of participants using each drug. ‘Not using’ = no drug use in the past month. **(C) Drug count by latent class.** Swarm plot of the number of drug types used (0–6) at TP2 in the two latent classes (Low-load, n=313; High-load, n=140); points are participants; diamonds and labels show class means. **(D) Drug use profiles by latent class.** Probability of using each drug given class membership in the two-class LCA solution; the dotted line marks 50%. **(E) PTSD symptoms by latent class.** PCL-5 scores at TP2 (n=354; Low-load, 240; High-load, 114). Points are participants; diamonds and labels show estimated means, and error bars their 95% confidence intervals (CI), adjusted for TP1 PCL-5, sex, age and time since the event; the bracket shows the regression contrast High vs. Low (** p<.01; Table S1). **(F) PTSD symptoms by number of drug types used.** Estimated means with 95% CI for 0–6 drugs (n=354; same adjustment as E). Brackets show regression coefficients versus 0 drugs (* p<.05, uncorrected for multiple comparisons; Table S1). **(G) PTSD symptoms by latent class, PCL-5≥33 only.** As in E, restricted to participants above the clinical cutoff at TP2 (n=227; Low-load, 157; High-load, 70); * p<.05 (Table S1).

**Table 1.** Demographic and clinical characteristics of the analytic sample at TP2 (N = 453)

|  | **Total N = 453** |  |
|---|---|---|
| **Characteristic** | **N or Mean** | **% or SD** |
| Sex |  |  |
| Men | 253 | 56 |
| Women | 200 | 44 |
| Age | 27.56 | 6.74 |
| Month since event at TP2 | 9.85 | 1.21 |
| Past Usage (prior to trauma exposure) |  |  |
| Abstinence / No use | 35 | 8.0 |
| Alcohol and/or cannabis | 29 | 6.6 |
| Poly-drug use | 345 | 78 |
| Single (not alcohol or cannabis) | 31 | 7.0 |
| PCL score at TP1 | 39.66 | 14.94 |
| PCL score at TP2 (8-12 Months) | 40.57 | 17.16 |
| PCL score at TP3 (17-24 Months) | 41.80 | 16.04 |

Values are n and % for categorical variables and mean and SD for continuous variables.

### Measures

Types of drug use were reported at TP1 for the traumatic event and at each follow-up assessment (TP2 and TP3), with a time frame of the past 30 days, via a multiple-choice questionnaire with an optional free-text field. For each drug used, participants also reported how many times they had used it in the past 30 days; these frequencies were used only to describe the latent classes (**Supplementary Methods**). Reported drugs were collapsed into six categories: alcohol, cannabis, ketamine, stimulants, MDMA, and psychedelics (**Supplementary Methods**). Binary indicators of use/nonuse were assigned to summarize patterns of co-use and derive latent classes.

Motives for drug use were reported using a checklist with optional free-text responses. After collapsing sparse categories, we retained five motive categories: avoidance, emotional regulation, self-medication (“to cure myself”), sleep, and recreational use.

PTSD symptom severity was assessed at TP1, TP2, and TP3 using the PTSD Checklist for DSM-5 (PCL-5) (10). Total scores were treated as a continuous variable for most analyses; we also used the established clinical cutoff (≥33) for specific subgroup analyses.

Demographic variables used as covariates were age at recruitment and sex. Time elapsed between October 7 and each outcome assessment was also included as a covariate. Whether participants had attended a party since October 7 and whether they had experienced an additional traumatic event since October 7 (yes/no) were compared between latent classes.

### Statistical analysis

Patterns of drug co-use, and motives, were visualized using UpSet plots. LCAs classified participants in three ways: (a) types of drugs used, (b) drug-use motives, and (c) a combined model incorporating both domains. Details of model selection and robustness analyses are in the **Supplementary Methods**. Because the drug-use classes were derived empirically, analyses of drug load and specific drug combinations were exploratory.

We then used multiple linear regression to examine associations between latent class membership (for each of three types of classification) and severity of PTSD symptoms. In additional analyses, we examined the total number of drugs used (0–6) as a categorical predictor. These regressions were repeated in subgroups above or below the cutoff for probable PTSD (PCL-5 ≥33). Associations of latent class membership with party attendance and additional traumatic-event exposure were evaluated with Pearson’s chi-squares. Longitudinal changes in PCL-5 scores between TP2 and TP3 were examined using linear mixed-effects models with all available PCL-5 observations, including fixed effects for timepoint, TP2 latent class, and their interaction, with participant-specific random intercepts. Regression models were adjusted for age, sex, and time elapsed since October 7. Primary analyses were additionally adjusted for baseline PTSD symptom severity (TP1 PCL-5 score). Mixed-effects models were adjusted for age and sex. Statistical analyses were conducted using R software. **Supplementary Table S1** summarizes statistical results for each figure.

## Results

### Sample

The analytic sample at TP2 comprised 453 trauma-exposed survivors who provided full drug-use data (**Table 1**). Data from TP1 regarding drug use during the event have been published (11).

### Polydrug use is frequent among mass trauma survivors

The UpSet plot (**Figure 1B**) shows that most participants (∼70%) reported using more than one drug in the month before TP2. Only 59 of 453 participants (13%) reported no drug use that month. The most common patterns were alcohol and cannabis (n=79) and cannabis only (n=37); other patterns involved combinations of three or more drugs, typically including psychedelics and MDMA.

Collapsing the polydrug combinations into a single drug while ignoring others indicated that people using MDMA or psychedelics (along with other drugs) showed lowest levels of PTSD symptoms: after adjustment, MDMA-users scored 4.28 points lower than non-MDMA-users (β=−4.28, Bonferroni-corrected p=.026) and psychedelic-users scored 3.90 points lower than non-psychedelic-users (β=−3.90, corrected p=.031).

In contrast, the differences for alcohol, cannabis, ketamine, and stimulants were not significantly different from non-users of these drugs (**Supplementary Figure S1**). Compared with non-user participants, only MDMA-users scored significantly lower (β=−6.62, corrected p=.017; **Supplementary Figure S2**). However, naturalistic co-use precluded tests for effects of any single drug. Therefore, LCA of group-level (load-based) comparisons were the primary analytic strategy, described below.

### High vs. low poly-drug use load

The first LCA, using the six binary drug indicators, identified a two-class solution, with classes differentiated primarily by number of drugs used rather than by which specific drugs were used: Low drug load class (n=313, mean=1.5 drug types) and High drug load class (n=140, mean=4.6 drug types; **Figure 1C**). The Low-load class was dominated by alcohol and cannabis use, each reported by over 50% of its members, with less use of other drug types. In the High-load class, all six drug groups exceeded the 50% threshold, with stimulants and psychedelics showing highest probabilities of use after alcohol and cannabis (**Figure 1D**).

### Higher drug load was associated with lower posttraumatic symptoms

Controlling for sex, age, time since event, and TP1 PCL-5 scores, survivors in the High-load class had lower PCL-5 scores at TP2 than those in the Low-load class (β=−3.71, 95% CI [−6.40, −1.01], p=.007, R²=.52; **Figure 1E**; statistics in **Supplementary Table S1**). A parallel model treating number of drugs used (0–6) as a 7-level factor (reference=0), with TP1 PCL-5 controlled, showed a similar pattern (overall effect of number of drugs, F(_6, 343_)=2.28, p=.036): PCL-5 scores did not differ from abstinence at 1–3 drugs and were lower at 4 or more drugs (4 drugs: β=−5.79, p=.017; 5 drugs: β=−7.26, p=.014; 6 drugs: β=−7.10, p=.024; uncorrected for multiple comparisons; **Figure 1F**; **Supplementary Table S1**).

This association did not simply reflect greater overall psychological health: when we restricted the sample to survivors with a current PCL-5 score above the clinical cutoff (≥33, n=227), the association had a similar direction and magnitude (β=−3.07, 95% CI [−5.65, −0.49], p=.020, with TP1 controlled; **Figure 1G**; **Supplementary Table S1**). Among survivors below the cutoff (n=127), the classes did not differ significantly (β=−1.13, 95% CI [−3.84, 1.58], p=.41; **Supplementary Table S1**). Thus, the drug-load association with PCL-5 is not fully explained by higher use among participants with fewer PTSD symptoms (for further control analyses see **Supplementary Results).**

### Motives for drug use

The UpSet plot of drug-use motives (**Figure 2A**) showed that recreational motives were the most commonly and uniquely endorsed (n=285 total endorsements; largest single unique group, n=84), followed by emotional management (n=230), sleep (n=193), avoidance (n=159), and self-medication (n=117). No dominant overlap pattern emerged; motives co-occurred within participants in a variety of combinations.

**Figure 2.**
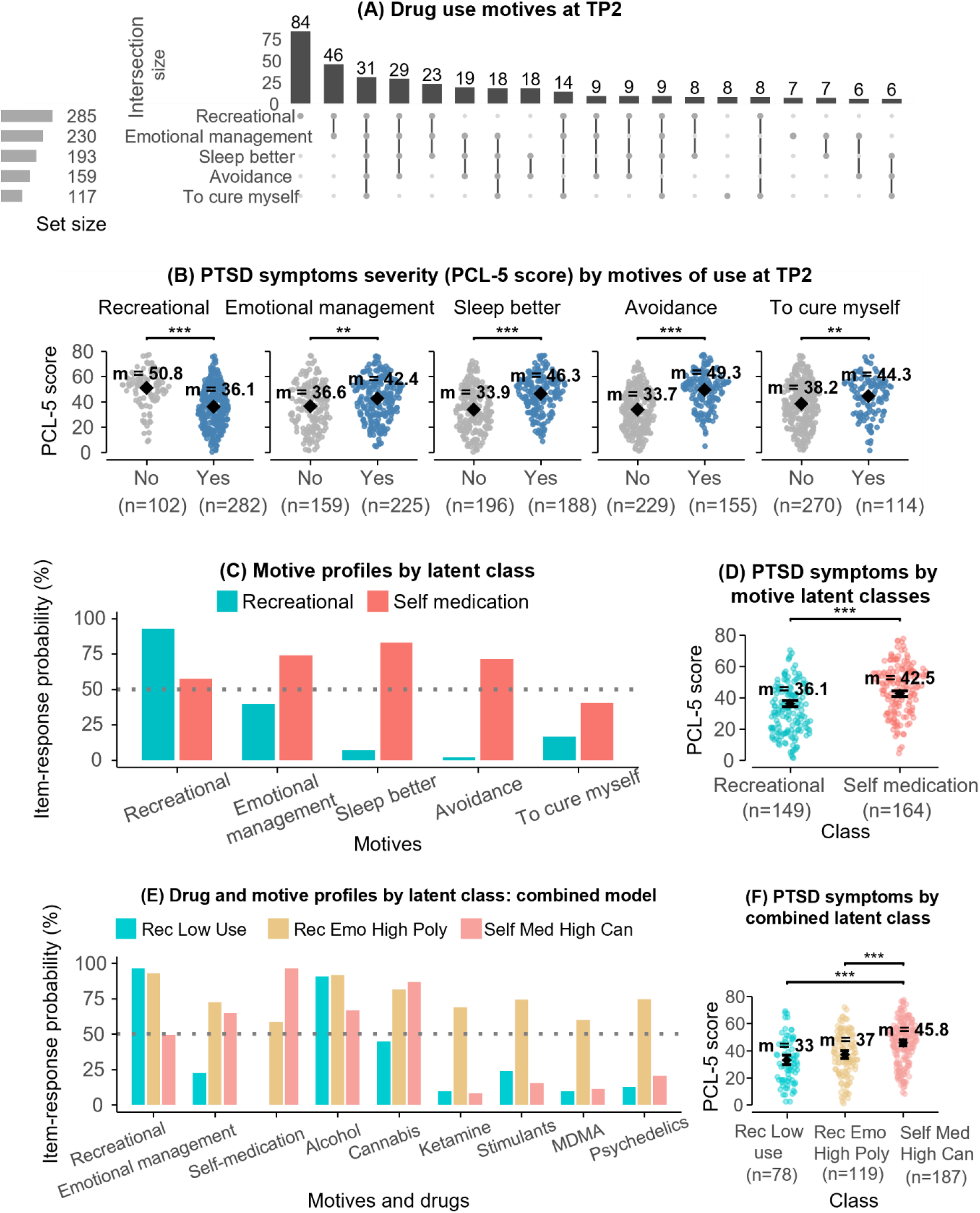
Drug use motives and PTSD symptoms. **(A) Drug use motives at TP2.** UpSet plot of endorsed motives (recreational, emotional management, sleep better, avoidance, to cure myself); layout as in Figure 1B. **(B) PCL-5 by endorsement of each motive.** For each motive, PCL-5 at TP2 in participants who did not (No) or did (Yes) endorse it (n=384). Points are participants; diamonds and labels show observed means; brackets show Welch t-tests, uncorrected for multiple comparisons (** p<.01, *** p<.001; Table S1). **(C) Motive profiles by latent class.** Probability of endorsing each motive given class in the two-class LCA (Recreational, Self-medication); the dotted line marks 50%. **(D) PTSD symptoms by motive-based latent class.** PCL-5 at TP2 (n=313; Recreational, 149; Self-medication, 164); diamonds and labels show estimated means, and error bars their 95% CI, adjusted for TP1 PCL-5, sex, age and time since the event; *** p<.001 (Table S1). **(E) Drug and motive profiles by latent class: combined model.** Probability of each item (three motives and six drugs) given class in the three-class combined LCA (Rec Low Use, Rec Emo High Poly, Self Med High Can). **(F) PTSD symptoms by combined latent class.** PCL-5 at TP2 (n=384; Rec Low Use, 78; Rec Emo High Poly, 119; Self Med High Can, 187); diamonds and labels show estimated means, and error bars their 95% CI, adjusted for sex, age and time since the event but not for TP1 PCL-5; brackets show Tukey-adjusted pairwise contrasts (*** p<.001; Table S1).

LCA on the five motive indicators identified two classes (**Figure 2C**): a “Recreational” class, predominantly endorsing fun/social use, and a “Self-medication” class, predominantly endorsing self-medication and/or emotional regulation. The motive classes differed mainly in cannabis use, which was about twice as frequent in the Self-medication class (median 30 vs. 15 times per month); alcohol (7 vs. 5) and stimulants (5 vs. 3) were also used somewhat more often, whereas psychedelics, ketamine and MDMA were used at similar frequencies (**Supplementary Figure S4**).

### Recreational motives were associated with lower posttraumatic symptoms

PCL-5 scores were higher in the Self-medication class (mean=42.5) than the Recreational class (mean=36.1) (β=6.37, 95% CI [3.60, 9.13], p<.001, controlling for sex, age, time since event, and baseline PCL-5 at TP1; **Figure 2D**; full model in **Supplementary Table S1**). An additional analysis tested each of the five motive categories separately (yes/no endorsement vs. PCL-5, simple uncorrected t-tests) rather than the LCA classes; results were broadly consistent with the LCA-based finding (**Figure 2B**).

### Combining drug type and motive identified three clinically meaningful classes

To integrate drug-type and motive information, we fit an LCA jointly on the six drug-type indicators and three collapsed motive indicators (self-medication, emotional regulation, recreational). This yielded a three-class solution (**Figure 2E**), labelled 1) “Self-medication and high cannabis use” (Self Med High Can; n=190); 2) “Recreation / emotional management and high polydrug use” (Rec Emo High Poly; n=122); and 3) “Recreation and low drug use” (Rec Low Use; n=79). The Rec Emo High Poly class endorsed use of all six drugs at high rates alongside elevated recreation and emotional-regulation motives; the Rec Low Use class endorsed almost exclusively alcohol and recreational motives, with essentially no self-medication (0%) and low rates of every other drug and motive.

### Recreational motives were again associated with lower posttraumatic symptoms

Group means adjusted for sex, age, and time since the event, but not for TP1 PCL-5 (**Figure 2F**), were higher in the Self Med High Can class (mean PCL-5=45.8) than in each of the other two recreational-use classes (Rec Emo High Poly, mean=37.0; β=-8.85, p<.001; Rec Low Use, mean=33.0; β=-12.85, p<.001). The two recreational-use classes did not differ significantly from one another (β=4.00, p=.197), despite considerably different patterns of drug use (See **Figure 2E**).

Some aspects of this pattern remained when we controlled for TP1 PCL-5, sex, age, and time-since-event (**Supplementary Table S1** and **Supplementary Figure S6A**): the Self Med High Can class remained significantly higher than the Rec Emo High Poly class (β=−4.74, Tukey-adjusted p=.007); the difference from Rec Low Use did not survive correction for multiple comparisons (β=−3.70, Tukey-adjusted p=.108; unadjusted p=.044). Again, the two recreational-type classes did not differ significantly from one another (β=-1.04, p=.847). However, adjusting for TP1 PCL-5 reversed the relative ranking of the two contrasts (**Supplementary Figure S6B**). Without TP1 adjustment (**Figure 2F**), the Rec Low Use class was further from Self Med High Can (β=−12.85) than the Rec Emo High Poly was (β=−8.85). With TP1 adjustment, Rec Emo High Poly showed the larger and more robust gap from Self Med High Can (β=−4.74, retaining ≈54% of its unadjusted magnitude), while Rec Low Use showed a smaller gap (β=−3.70, retaining only ≈29%).

In other words, most of the Rec Low Use class’s apparent PCL-5 advantage in the raw comparison (**Figure 2F**) appeared to reflect pre-existing (TP1) differences in PTSD scores rather than correlates of their current drug-use pattern. In contrast, the Rec Emo High Poly class’s difference from the Self Med High Can class, held up independent of TP1 PTSD severity. We therefore report both models: **Figure 2F** for the group separation without TP1 adjustment and **Supplementary Figure S6** for the severity-adjusted comparison.

### Longitudinal follow-up

Of the 453 participants with TP2 drug-use data, 302 (67%) also had drug-use data at TP3 (17-24 months post-attack). The TP3 UpSet plot (**Figure 3A**) shows the same overall polydrug-use-dominant, alcohol/cannabis-led hierarchy of drug combinations observed at TP2.

**Figure 3.**
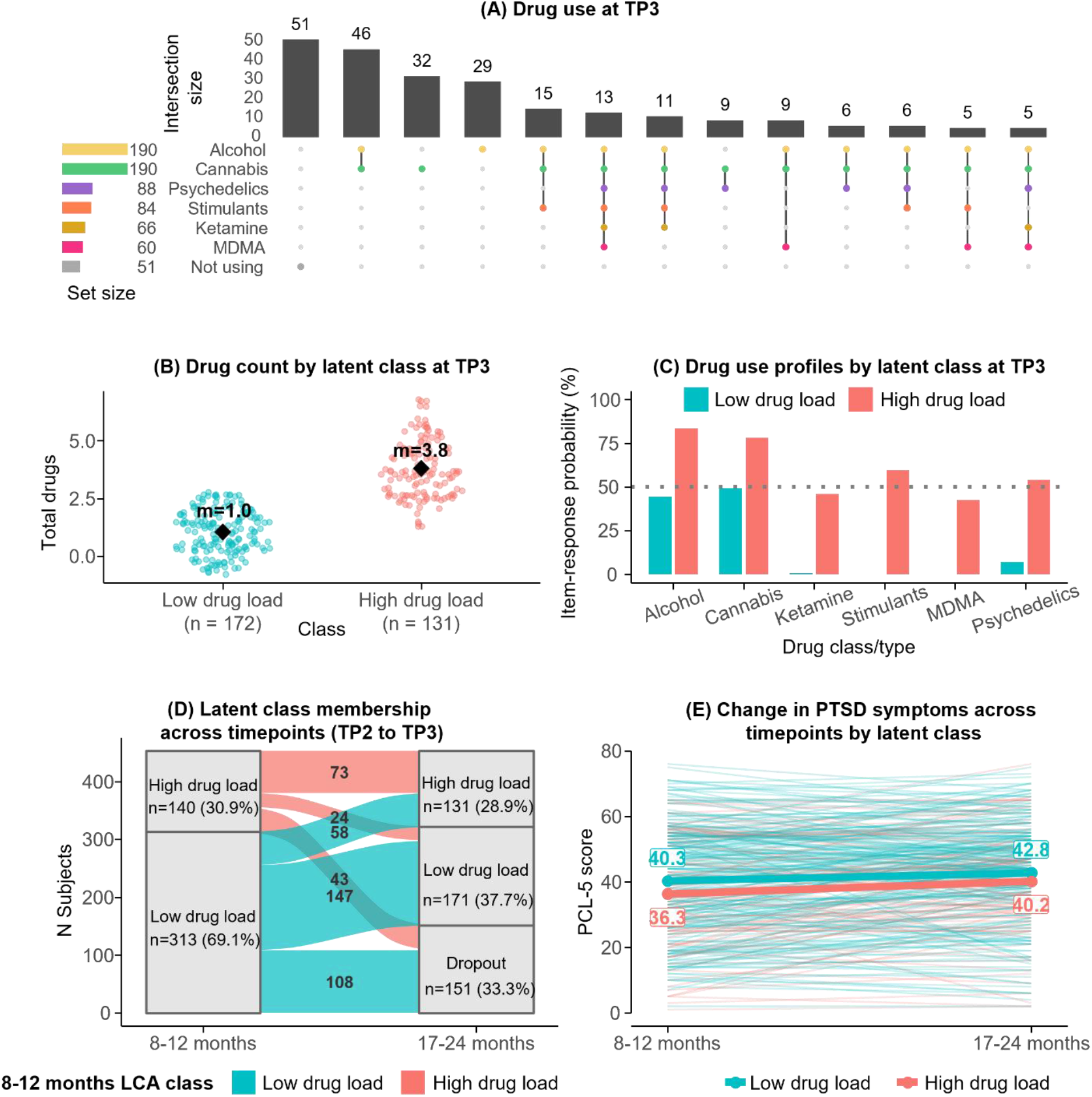
Drug use patterns and PTSD symptoms at TP2 and TP3. **(A) Drug use at TP3.** UpSet plot of the drug combinations reported at TP3 among the 303 participants with drug-use data at TP3 (combinations with at least 5 participants are shown; layout as in Figure 1B). **(B) Drug count by latent class at TP3.** Swarm plot of the number of drug types used at TP3 in the two classes of the TP3 LCA (Low-load, n=172; High-load, n=131); diamonds and labels show class means. n=172 includes one participant classified at TP3 who had no TP2 class and is therefore not in panel D (Low-load at TP3 = 171). **(C) Drug use profiles by latent class at TP3.** Probability of using each drug given class membership in the TP3 LCA; the dotted line marks 50%. **(D) Latent class membership across timepoints (TP2 to TP3).** Alluvial diagram of transitions between TP2 and TP3 classes for the 453 participants classified at TP2, including dropout (n=151); 302 participants had a class at both timepoints. The blocks show class sizes (N, % of the timepoint) and the flows show transition N. **(E) Change in PTSD symptoms across timepoints by latent class.** PCL-5 at TP2 and TP3 for participants with a valid score at both timepoints (n=292; Low-load, 196; High-load, 96), colored by TP2 class. Thin lines are participants; bold lines and labels show observed class means. The mixed model (742 observations from 450 participants) showed main effects of timepoint (p<.001) and class (p=.032) and no interaction (p=.333; Table S1).

Re-fitting our LCA onto the TP3 drug indicators (303 participants with TP3 drug-use data, 302 of whom were also classified at TP2; the remaining participant had only a partial TP2 response), we again identified two classes: the same Low-load/High-load structure identified at TP2 (**Figure 3B-C**), suggesting that this was a stable feature of the sample. Class membership was moderately stable across time (**Figure 3D**): among participants retained at both timepoints, 75% of the TP2 High-load class remained High-load at TP3 (73/97), and 72% of the TP2 Low-load class remained Low-load (147/205); the remaining participants transitioned across the load boundary in both directions (25% High→Low, 28% Low→High). Attrition between TP2 and TP3 was similar across classes (30.7% of the High-load class vs. 34.5% of the Low-load class dropped out; χ²(1)=0.47, p=.49; **Supplementary Table S1**), so differential dropout is unlikely to account for the transitions.

Finally, a linear mixed model testing PCL-5 change from TP2 to TP3 as a function of the TP2 drug-load class (**Figure 3E**) showed a significant main effect of time (PCL-5 score increased on average by 2.67 points across timepoints, F_(1, 312.4)_=13.75, p<.001) and of load class (High-load lower than Low-load by 3.48 points on average across timepoints, F_(1, 447.3)_=4.65, p=.032), but no interaction (F_(1, 312.7)_=0.94, p=.333): both classes showed a similar, modest worsening of posttraumatic symptoms across the second year after the event.

## Discussion

In previously reported results from this sample of mass-trauma survivors, we found that being acutely under the influence of MDMA or psychedelics *during* the event was perceived as helpful for trauma processing. For MDMA, this was accompanied by lower mental distress, better sleep, and more social interactions in the four months after the event (11). In contrast, cannabis and alcohol use were associated with worse symptoms (11). Here, we report follow-up data on ongoing drug use and its relationship to PCL-5 scores up to two years after that traumatic event.

We report three main findings. First, at TP2 (8-12 months), ongoing polydrug use was common, and use of more drug types (including, though not limited to, MDMA and psychedelics) was unexpectedly associated with lower PCL-5 scores compared with use of fewer drug types (mostly, but not only, alcohol and cannabis). Additionally, the association between higher polydrug use and lower PTSD symptom severity persisted after adjustment for baseline (TP1) PTSD symptoms, and among participants whose PCL-5 scores exceeded the cutoff for PTSD.

Second, motives for drug use were associated with PTSD symptom severity regardless of the number of drug types used: participants reporting self-medication motives had higher PCL-5 scores than those reporting recreational motives.

Third, the association between lower PCL-5 scores and the use of more drug types was largely preserved longitudinally at TP3 (17-24 months), although PTSD symptom severity increased modestly over time regardless of the number of drug types used.

### Drug use after mass trauma

Increased drug use after mass trauma is well documented. Several studies reported increased drug use among Manhattan residents after the World Trade Center attack (13, 14), and meta-analyses show increases after other terrorist attacks (15, 16).

Two surveys of Israeli adults conducted about one month after October 7, 2023 found comparable increases. In one, 34% reported increased use of at least one drug, most often tobacco, sedatives, pain medication, and alcohol (only 5.5% reported increased cannabis use), and greater trauma exposure predicted increased use of sedatives but not other drugs (17). In the other study, 19% reported increased drug use, most often alcohol, followed by sedatives and cannabis; increased use was more likely after direct exposure (OR=5.75) and with greater psychological distress (OR=1.80) (18).

Increased drug use after trauma is often viewed as a maladaptive coping strategy, a form of “self-medication” in which people use drugs that temporarily reduce specific posttraumatic symptoms (18–20). The effect of such self-medication remains unclear. In a large epidemiological study, self-medication of PTSD symptoms was associated with greater psychiatric comorbidity and more suicide attempts (3). Although causality is difficult to establish, drug use after trauma is generally considered detrimental to clinical outcomes (20, 21).

Unexpectedly, our data do not support this association. In fact, Nova survivors who used more drug types had lower PTSD symptom severity than those who used fewer, and this association persisted after adjustment for baseline symptoms and over the second year after the attack.

Several differences between our study and previous work may explain this discrepancy. First, prior research has largely examined the relationship between PTSD and SUDs, typically in clinical, help-seeking, or veteran cohorts in which problematic drug use was the defining feature (20, 22–25). In contrast, our cohort comprised civilian survivors of a mass terror attack at a music festival, most of whom had a history of recreational drug use before the attack and many of whom developed clinically significant PTSD symptoms after it. Increased drug use after trauma does not necessarily indicate disorder: in a meta-analysis of adults exposed to terrorist attacks, 6% increased their alcohol consumption, but the prevalence of alcohol use disorder was no higher than in the general population (16).

Second, prior work has focused mainly on alcohol, cannabis, sedatives, and opioids (17, 18), and has often examined a single drug in isolation (e.g. 21, 26, 27). We characterized naturalistic polydrug use patterns, which were complex and varied across individuals. Our “high load” class was distinguished by use of MDMA, psychedelics, and ketamine, drug types under clinical investigation as medications for PTSD (6, 8). In single-drug analyses, users of MDMA and psychedelics showed the largest symptom differences relative to non-users. However, because these drugs were almost always used as part of a complex polydrug pattern, the association cannot be attributed to any single drug or drug class. Motives for use appear to matter: drug load and self-medication motives were associated with symptom severity in opposite directions.

## Limitations

Our observational study has several limitations. First, we did not assess prior or emergent symptoms of SUDs, nor perceptions of benefit from ongoing drug use. Thus, we cannot draw direct conclusions about the benefits or harms of the ongoing drug use reported.

Second, class differences in PCL-5 scores, even when statistically significant, were small in absolute terms. The sample as a whole had exceedingly high PCL-5 scores, reflecting severe persisting PTSD symptoms, more than has been seen in other longitudinal studies of trauma survivors, including mass-trauma survivors (28, 29).

Third, drug load may partly reflect social re-engagement rather than successful self-treatment. High-load participants were more likely than Low-load participants to have attended a party since October 7. For festival survivors, returning to parties and their associated drug use may mark a return to pre-trauma routines, which may be more accessible to those with fewer symptoms.

Our findings do not tell a simple story of either benefit or harm from ongoing drug use: there is no clear evidence that drug use in our participants exacerbated their PTSD symptoms, but also no clear evidence that drug use resolved them. We cannot answer the central counterfactual question: how many participants’ PCL-5 scores would now be either lower or higher if they had *not* used their self-selected combinations of drugs at self-selected frequencies? The current data might reflect a combination of desirable and undesirable outcomes for each pattern of drug use and each class of motives.

## Conclusions and clinical implications

Polydrug use was common among Nova festival survivors up to two years after the attack. Higher drug load, involving more frequent use of MDMA and psychedelics, was associated with lower PTSD symptom severity, whereas self-medication motives, accompanied by near-daily cannabis use, were associated with higher severity. The cannabis finding is consistent with our earlier report that cannabis use during the attack predicted worse psychological outcomes (11), and with prospective data from US veterans (21, 30). This, together with the well-documented risks of alcohol in PTSD (20, 22), suggest that clinicians should ask trauma survivors not only which drugs they use but why.

Finally, non-addictive drug use has been proposed to be instrumental, serving goals such as improved social interaction or coping with stress (31). Our findings fit this framework: motives for use, not only amount or type of drug, were associated with posttraumatic symptoms. The MDMA and psychedelic findings are compatible with, but do not demonstrate, a therapeutic effect, and do not support unsupervised use as treatment. Although long-term use of addictive drugs can be harmful, recreational use of some drug classes, such as MDMA and psychedelics, may be associated with better outcomes in some people under some circumstances, including after severe trauma.

## Data Availability

Due to the sensitive nature of the study population and privacy considerations, individual participant data are not publicly available. De-identified data may be made available from the corresponding author upon reasonable request and subject to institutional ethics review.

## Acknowledgments

We are deeply grateful to the survivors and all participants for their time and trust during an exceptionally difficult period in their lives; their generosity made this work possible. We also thank SafeHeart for facilitating survivor outreach and support. This writing of this manuscript was supported [in part] by the Intramural Research Program of the National Institutes of Health (NIH, DE, YS). The contributions of the NIH author(s) are considered Works of the United States Government. The findings and conclusions presented in this paper are those of the author(s) and do not necessarily reflect the views of the NIH or the U.S. Department of Health and Human Services.

