## Supplementary Materials for "Polydrug use and recreational motives are associated with lower posttraumatic symptoms in survivors of a mass trauma event"

### Supplementary Methods

**Survey design:** At TP1 (1-6 months after the Nova festival), participants provided demographic information, including age and sex assigned at birth, and completed questionnaires assessing trauma exposure and drug use during the Nova festival preceding the traumatic event. Drug use since the attack, at TP2 (8-12 months) and TP3 (17-24 months), was assessed within a dedicated survey block. Participants first answered a single gating item: "Did you consume psychoactive drugs in the last month (including alcohol and cannabis)?" (Yes/No); participants answering "No" skipped the remainder of the block and were coded as reporting no drug use for that period. Participants who responded affirmatively were presented with a check-all-that-apply drug checklist ("Please mark if you took any of the drugs from the following list in the last month"). They were then asked to indicate their reasons for drug use during the previous month ("For what purpose did you take the drugs you mentioned in the last month?"; multiple selections permitted).

PTSD symptom severity was assessed at all three time points using the PTSD Checklist for DSM-5 (PCL-5), a 20-item self-report measure of current PTSD symptoms. Total scores range from 0 to 80, with scores ≥33 indicating probable PTSD diagnosis. Questionnaires were administered in Hebrew; English translations are provided in this manuscript for reporting purposes. The survey was administered through the Qualtrics platform, which enabled secure, anonymous, and confidential data collection.

**Drug categorization:** The drug checklist presented to participants included: cannabis, MDMA, ketamine, LSD, mushrooms, cocaine, DMT, 4-methylmethcathinone, ecstasy, 2C-B, alcohol, and a free-text "Other (please specify)" option. Drug responses were collapsed into six analytical categories: Cannabis, Alcohol, and Ketamine were each retained as their own category; MDMA and ecstasy were combined into the MDMA category; cocaine, 4-methylmethcathinone, and amphetamines were combined into the Stimulant category; and LSD, psilocybin, DMT and 2C-B were combined into the Psychedelics category.

Free-text responses were reviewed and manually recoded: entries pharmacologically consistent with one of the six established categories were folded into that category; prescription psychiatric medications (e.g., fluoxetine, lamotrigine, venlafaxine) were coded as a separate "Prescription" group; drugs not matching any established category (e.g., sleeping pills, GHB) were coded as "Other"; and responses too ambiguous to classify (e.g., explicit refusals to specify) were coded as "Unknown." The “Prescription”, “Other” and “Unknown” were excluded from the main analyses. Drug categories were defined in consultation with experts in psychoactive drugs, harm reduction practitioners, and survivors of the Nova festival attack.

**Categorization of motives for drug use:** Participants selected one or more reasons for drug use from the following predefined options: "To feel good / for a feeling of euphoria," "To sleep better," "To avoid memories / to forget," "To avoid dreams," "To heal myself," "For fun / in a social setting," and a free-text option ("Other reason, please specify"). Responses were collapsed into five motive categories for analyses. "For fun / in a social setting" was retained as the Recreational category; "To sleep better" as the Sleep category; and "To heal myself" as the Self-medication category. "To avoid memories / to forget" and "To avoid dreams" were combined into the Avoidance category, whereas "To feel good / for a feeling of euphoria" was classified as Emotional Regulation. Free-text responses were reviewed and manually re-coded. Most were assigned to one of the five predefined categories (e.g., "to relax" and "to handle a bad day" → emotional regulation; "to disconnect from reality" → avoidance).

A smaller number were initially assigned to additional intermediate categories not included in the final analyses, including spiritual (e.g., "to deepen my consciousness"), medical/physical state (e.g., chronic pain, fibromyalgia, epilepsy), cognitive/physical function (e.g., "to be able to focus/study"), and habit/addiction. In the combined modelling of drugs and motives we collapsed all three motives categories involving PTSD symptoms (Avoidance, Sleep and 'Healing myself') into the Self-Medication category.

**Individual drugs and PTSD symptoms:** To examine whether specific drugs were associated with PTSD symptoms irrespective of co-use patterns, we ran separate regression models for each of the six drug categories, collapsing polydrug combinations into a single drug while ignoring the others. In the first analysis, PCL-5 at TP2 was regressed on use of the drug in the month before TP2 (yes/no), regardless of the use of other drugs. In the second analysis, users of each drug were compared with participants reporting abstinence from all drugs (n=59). All models included sex, age, months since October 7 and TP1 PCL-5 as covariates. p-values for the drug coefficient were Bonferroni-corrected for the six drugs.

**Latent class analysis:** Latent class analyses were conducted using the poLCA package (SR1). Separate LCAs were fitted to drug-use indicators, drug-use motives, and the combined indicator set. Models specifying one to five classes were evaluated using the Bayesian Information Criterion (BIC). Each model was estimated using multiple random expectation-maximization (EM) starts (50 random starts for the drug-type and motive models; 10 for the combined model). The optimal solutions consisted of two drug-use classes, two motive classes, and three combined drug-motive classes.

**Frequency of drug use:** For each drug that participants reported using in the 30 days before the assessment, they also stated on how many of those days they had used it. Numeric responses were retained (including decimals); non-numeric responses (e.g., "unknown") were treated as missing; and participants who did not use a drug were not included in that drug's distribution ("users only"). Frequency was summarized descriptively (times used per month; mean, SD and median).

For the whole sample, we plotted the number of users at each reported frequency. To describe how the latent classes (drug-load, motive and combined classes) differ in frequency of use, we plotted for each drug the percentage of each class's users at each frequency, which accounts for the different class sizes. Because frequency was self-reported in free text and strongly right-skewed, with heaping at round values (e.g., 30 times per month for cannabis), we did not model it, and it was not used to define the classes or as a covariate. No statistical tests were performed.

**Regression analyses:** Associations between latent class and PCL-5 at TP2 were tested with multiple linear regression. All regression models included sex, age at recruitment, and the months elapsed between October 7, 2023 and the assessment of the outcome as covariates. Primary models additionally included the TP1 PCL-5 score ("TP1-adjusted"); the combined-class model is reported both without and with this adjustment. The total number of drugs used (0–6) was entered as a seven-level categorical predictor with abstinence (0 drugs) as the reference; each level was compared with the reference using the regression coefficients, without correction for multiple comparisons. For the three-class combined model, pairwise comparisons used Tukey-adjusted estimated marginal means (emmeans package; SR2). Comparisons between two groups required no correction. The regression analyses were repeated in the subgroups with PCL-5≥33 and <33 at TP2; because these subgroups are defined by the outcome, their estimates should be interpreted descriptively.

**Welch t-tests:** Differences in PCL-5 at TP2 between participants who did and did not endorse each of the five motives (Figure 2B) were tested with two-sided Welch t-tests, without covariates and without correction for multiple comparisons.

**Chi-square tests:** Associations of the drug-load classes with party attendance since October 7 and with exposure to an additional traumatic event since October 7 (yes/no items; participants without a valid answer were excluded, n=445 for each) were tested with Pearson's chi-square tests with Yates' continuity correction, without covariates. The same test was used to compare dropout between TP2 and TP3 across TP2 classes (n=453).

**Bayes factors:** To complement the frequentist contrasts, we computed Bayes factors (BF10) for each pairwise comparison between classes with the BayesFactor R package (version 0.9.12-4.8; SR3). For each pair, a linear model was fitted to the participants in the two classes being compared, and BF10 was defined as the evidence for a model that includes the class variable relative to a model containing the covariates only (sex, age at recruitment, months since October 7 and, in models marked "with baseline", the baseline score of the same outcome). The covariates were included in both models. We used the package's default JZS priors: a Cauchy prior with scale r=0.707 on the standardized difference between the two classes (and on categorical covariates) and r=0.354 on continuous covariates (SR4).

The test is two-sided (BF10>1 favors a difference between classes; BF10<1 favors no difference) and, like the regression models, assumes normally distributed residuals with equal variance. For two-class predictors, the Bayes factor uses all participants and corresponds to the regression contrast. For predictors with more than two levels (the three-class combined model and the number-of-drugs analysis), each Bayes factor is based on the two groups being compared, whereas the reported differences and p-values come from the model containing all groups. For the combined model these are Tukey-adjusted pairwise contrasts of estimated marginal means (emmeans package; SR2); for the number of drugs they are regression coefficients versus 0 drugs, not corrected for multiple comparisons.

Bayes factors are not adjusted for multiple comparisons; the two are therefore different tests and should not be compared directly. Bayes factors were estimated by Monte Carlo integration (100,000 iterations; fixed random seed, 2024), with a numerical error of roughly 1–2% in our checks. Evidence was labelled (SR5): BF10>3, >10, >30 and >100 indicate moderate, strong, very strong and extreme evidence for a difference, and BF10<1/3, <1/10, <1/30 and <1/100 indicate the corresponding evidence for no difference; values between 1/3 and 3 are anecdotal.

**Longitudinal analyses:** Longitudinal changes in PTSD symptom severity from TP2 to TP3 were examined using linear mixed-effects models with participant-specific random intercepts (lme4 package; SR6). Fixed effects were evaluated using Type III tests with the Satterthwaite approximation. The model included timepoint, TP2 drug-load class, their interaction, sex and age; months since October 7 was not included because it is determined by timepoint. All available PCL-5 observations were used.

**Software:** All analyses were performed in R version 4.6.0 (emmeans 2.0.4, BayesFactor 0.9.12-4.8, lme4 2.0.1, lmerTest 3.2.1).

### Supplementary Results

**Supplementary Table S1** summarizes the statistics for each panel of Figures 1–3 and for the analyses reported only in the main text: the test statistic, p-value, effect estimate with 95% confidence interval, and Bayes factor where computed.

**Individual drugs and PTSD symptoms: Supplementary Figure S1** compares users and non-users of each drug, and **Supplementary Figure S2** compares users of each drug with abstinent participants. Users of every drug had lower PCL-5 scores than non-users after adjustment ($\beta=-1.77$ to $-4.28$), but only MDMA ($\beta=-4.28$; unadjusted means 35.9 vs. 42.0) and psychedelics ($\beta=-3.90$;

37.3 vs. 42.0) remained significant after Bonferroni correction for the six drugs (corrected p=.026 and .031, respectively). Compared with abstinent participants (unadjusted mean PCL-5=43.8), all six differences were in the same direction (β=−3.81 to −6.62), but only MDMA-users differed significantly (β=−6.62, corrected p=.017); psychedelic-users showed the second-largest difference without reaching significance after correction (β=−5.40, corrected p=.082). These analyses should be interpreted with caution. The categories overlap, since most users of one drug also used others, so each coefficient reflects use of that drug in the context of the participant's usual co-use and cannot be attributed to the drug itself. MDMA and psychedelics were far more prevalent in the High-load class than in the Low-load class (Figure 1D), so these associations may partly reflect overall drug load. The abstinent group is small (n=59 with TP2 PCL-5), which limits power. The figures show observed means and group sizes for all participants with a PCL-5 score at TP2, whereas the regression models include only participants with TP1 PCL-5 data (n=354 for users vs. non-users; n=133–298 for comparisons with abstinent participants), so β differs from the raw mean differences.

**Low- vs. High-load classes:** In a complementary analysis, we tested whether the Low- vs. High-load classes differed on two variables collected at TP2: party attendance and exposure to additional traumatic events.

Party attendance since October 7 was more common in the High-load class (96%) than in the Low-load class (80%; $\chi^2_{(1)} = 17.56$, $p < .001$). In contrast, self-reported exposure to additional traumatic events since October 7 did not differ between groups ($\chi^2_{(1)} = 1.12$, $p = .29$; ~44–50% in both classes), indicating that differences in drug use were not simply driven by differential exposure to subsequent adverse events.

**Frequency of use:** Whole sample: Among users who reported how often they used each drug, frequency of use differed markedly by drug (**Supplementary Figure S7**). Cannabis was typically used daily (median 30 times per month; n=295), alcohol had a more even distribution (median 6; n=307), and psychedelics, stimulants, ketamine and MDMA were used infrequently (median 2–3; right-skewed with a mode at 1–2 times per month; n=106–143). Drug-load classes **(Supplementary Figure S3)**: The classes differed in frequency of use as well as in the number of drugs used. High-load users used alcohol (median 8 vs. 5 times per month; mean 10.3 vs. 8.0), psychedelics (3 vs. 1; 4.4 vs. 2.9), stimulants (4 vs. 2; 6.3 vs. 5.0) and MDMA (2 vs. 1; 2.9 vs. 1.4) more often than Low-load users.

Cannabis frequency did not differ (median 30 in both classes; mean 23.3 vs. 23.5). Ketamine had a higher median in the High-load class (4 vs. 2) but similar means (5.8 vs. 5.9), reflecting a few Low-load users reporting 10–30 uses among only 17 Low-load ketamine users. Motive classes **(Supplementary Figure S4)**: The classes differed mainly in cannabis use, which was about twice as frequent among Self-medication users (median 30 vs. 15; mean 27.5 vs. 16.2). Alcohol (7 vs. 5; 10.2 vs. 7.5) and stimulants (5 vs. 3; 7.0 vs. 5.0) were also used somewhat more often, whereas psychedelics (median 2 in both classes), ketamine (4 vs. 3) and MDMA (median 2 in both) were used at similar frequencies.

Combined classes **(Supplementary Figure S5)**: The classes differed most in cannabis and alcohol use, which were least frequent in Rec Low Use (cannabis median 7 vs. 30 in both other classes, mean 11.4 vs. 23.3 and 26.0; alcohol 5 vs. 8 and 7, mean 6.2 vs. 10.4 and 9.1), and in psychedelics and MDMA use, which were most frequent in Rec Emo High Poly (psychedelics median 3 vs. 1 in both other classes, mean 4.6 vs. 1.6 and 3.1; MDMA 2 vs. 1 in both, mean 3.2 vs. 1.2 and 1.5). The two high-cannabis classes used cannabis equally often (median 30) and did not differ clearly in stimulant use (median 4 vs. 3; mean 6.4 vs. 5.9) or ketamine use (4 vs. 2.5; mean 5.6 vs. 6.8). Rec Low Use had only 5–18 users of psychedelics, stimulants, ketamine and MDMA, so comparisons involving this class for those drugs are descriptive only. These comparisons are descriptive: no tests were performed, frequency is self-reported and heaped at round values, and several class-by-drug groups are small.

SR5. Lee MD, Wagenmakers E-J: Bayesian Cognitive Modeling: A Practical Course. Cambridge, UK, Cambridge University Press, 2013

SR6. Bates D, Mächler M, Bolker B, et al.: Fitting linear mixed-effects models using lme4. J Stat Softw 2015; 67:1–48

**Table S1.** Statistical summary for the figure panels that include inferential tests (Figures 1–3).

| Panel | Data | Test | Effect | Statistic | p | Estimate [95% CI] | BF10 |
|---|---|---|---|---|---|---|---|
| **Figure 1** | | | | | | | |
| 1E | PCL-5 at TP2 by drug load class (n = 354) | Linear regression, TP1-adjusted | Class: High − Low | t(348) = −2.71 | .007 | −3.71 [−6.40, −1.01] | 4.0 |
| 1F | PCL-5 at TP2 by number of drugs used (0–6; n = 354) | Linear regression, TP1-adjusted; number of drugs as 7-level factor | Number of drugs (overall) | F(6, 343) = 2.28 | .036 | n/a | n/a |
| | | Coefficients vs 0 drugs, uncorrected | 1 vs 0 | t(343) = −0.54 | .59 | −1.31 [−6.09, 3.48] | 0.24 |
| | | | 2 vs 0 | t(343) = −0.83 | .41 | −1.80 [−6.06, 2.47] | 0.21 |
| | | | 3 vs 0 | t(343) = −1.07 | .29 | −2.57 [−7.29, 2.15] | 0.37 |
| | | | 4 vs 0 | t(343) = −2.39 | .017 | −5.79 [−10.56, −1.03] | 1.5 |
| | | | 5 vs 0 | t(343) = −2.46 | .014 | −7.26 [−13.05, −1.46] | 1.8 |
| | | | 6 vs 0 | t(343) = −2.27 | .024 | −7.10 [−13.25, −0.94] | 1.7 |
| 1G | PCL-5 at TP2 among participants with PCL-5 ≥ 33 (n = 227) | Linear regression, TP1-adjusted | Class: High − Low | t(221) = −2.35 | .020 | −3.07 [−5.65, −0.49] | 2.1 |
| *Text only* | *PCL-5 at TP2 among participants with PCL-5 < 33 (n = 127)* | Linear regression, TP1-adjusted | Class: High − Low | t(121) = −0.83 | .41 | −1.13 [−3.84, 1.58] | 0.27 |
| *Text only* | *Party attendance since October 7 by class (n = 445)* | Pearson $\chi^2$ test, Yates' continuity correction | Class × party attendance | $\chi^2(1)$ = 17.56 | <.001 | n/a | n/a |
| *Text only* | *Additional traumatic event since October 7 by class (n = 445)* | Pearson $\chi^2$ test, Yates' continuity correction | Class × additional event | $\chi^2(1)$ = 1.12 | .29 | n/a | n/a |
| **Figure 2** | | | | | | | |
| 2B | PCL-5 at TP2 by endorsement of each motive, yes/no (n = 384 per test) | Welch t-test, uncorrected | Recreational: Yes − No | t(201.3) = −8.41 | <.001 | −14.64 [−18.08, −11.21] | n/a |
| | | | Emotional management: Yes − No | t(321.0) = 3.26 | .001 | 5.84 [2.31, 9.36] | n/a |

| Panel | Data | Test | Effect | Statistic | p | Estimate [95% CI] | BF10 |
|---|---|---|---|---|---|---|---|
| | | | Sleep better: Yes − No | t(379.1) = 7.56 | <.001 | 12.39 [9.17, 15.61] | n/a |
| | | | Avoidance: Yes − No | t(352.0) = 9.87 | <.001 | 15.56 [12.46, 18.66] | n/a |
| | | | To cure myself: Yes − No | t(231.0) = 3.30 | .001 | 6.06 [2.45, 9.68] | n/a |
| 2D | PCL-5 at TP2 by motive-based class (n = 313) | Linear regression, TP1-adjusted | Class: Self medication − Recreational | t(307) = 4.53 | <.001 | 6.37 [3.60, 9.13] | >100 |
| 2F | PCL-5 at TP2 by combined class (n = 384) | Linear regression, no TP1 adjustment; Tukey-adjusted p and CI | Self Med High Can − Rec Low Use | t(378) = 5.98 | <.001 | 12.85 [7.79, 17.90] | >100 |
| | | | Self Med High Can − Rec Emo High Poly | t(378) = 4.74 | <.001 | 8.85 [4.45, 13.25] | >100 |
| | | | Rec Emo High Poly − Rec Low Use | t(378) = 1.73 | .197 | 4.00 [−1.46, 9.45] | 0.55 |
| S6 | *PCL-5 at TP2 by combined class, TP1-adjusted (n = 313)* | Linear regression, TP1-adjusted; Tukey-adjusted p and CI | Self Med High Can − Rec Emo High Poly | t(306) = 3.06 | .007 | 4.74 [1.09, 8.40] | 14 |
| | | | Self Med High Can − Rec Low Use | t(306) = 2.02 | .108 | 3.70 [−0.61, 8.01] | 1.7 |
| | | | Rec Emo High Poly − Rec Low Use | t(306) = −0.55 | .847 | −1.04 [−5.52, 3.44] | 0.22 |
| **Figure 3** | | | | | | | |
| 3D | Dropout between TP2 and TP3 by TP2 class (n = 453) | Pearson $\chi^2$ test, Yates' continuity correction | Class × dropout | $\chi^2(1) = 0.47$ | .49 | n/a | n/a |
| 3E | PCL-5 at TP2 and TP3 (742 observations, 450 participants) | Linear mixed model, random intercept; sex- and age-adjusted | Timepoint (TP3 − TP2) | F(1, 312.4) = 13.75 | <.001 | 2.67 [1.25, 4.08] | n/a |
| | | Satterthwaite F-tests | Class (High − Low, averaged over timepoints) | F(1, 447.3) = 4.65 | .032 | −3.48 [−6.66, −0.31] | n/a |
| | | | Timepoint × class | F(1, 312.7) = 0.94 | .333 | n/a | n/a |

Software: R 4.6.0; linear models (lm), estimated marginal means and pairwise contrasts (emmeans 2.0.4), Welch t-tests (t.test), chi-square tests (chisq.test), linear mixed models (lme4 2.0.1; lmerTest 3.2.1, Satterthwaite degrees of freedom), Bayes factors (BayesFactor 0.9.12-4.8).

Estimates are the difference between the groups named in the Effect column (first minus second); 95% confidence intervals are unadjusted, except for the three-class combined model, where they are Tukey-adjusted like the p-values. All p-values are two-sided. Regression models were adjusted for sex, age and months since October 7; "TP1-adjusted" models additionally include TP1 PCL-5. The mixed model was adjusted for sex and age. Welch t-tests and chi-square tests were unadjusted.

Multiple comparisons: p-values for the three-class combined model are Tukey-adjusted. Coefficients for number of drugs (1F) are tested against 0 drugs and are not corrected for multiple comparisons; Welch t-tests (2B) are not corrected. Comparisons between two groups need no correction.

BF10: Bayes factor for the model with versus without the predictor, with the covariates in both models (JZS prior; Cauchy scale r = 0.707 for the group difference and r = 0.354 for continuous covariates; two-sided). For predictors with more than two levels, each Bayes factor is based on the two groups compared and is not corrected for multiple comparisons. BF10 > 3, > 10, > 30 and > 100 indicate moderate, strong, very strong and extreme evidence for a difference; BF10 < 1/3 indicates moderate evidence for no difference. n/a = not computed.

"Text only" rows report analyses described in the Results that are not plotted. Panels without inferential statistics (1A–D, 2A, 2C, 2E, 3A–C) are not listed. PCL-5, PTSD Checklist for DSM-5; TP, timepoint.

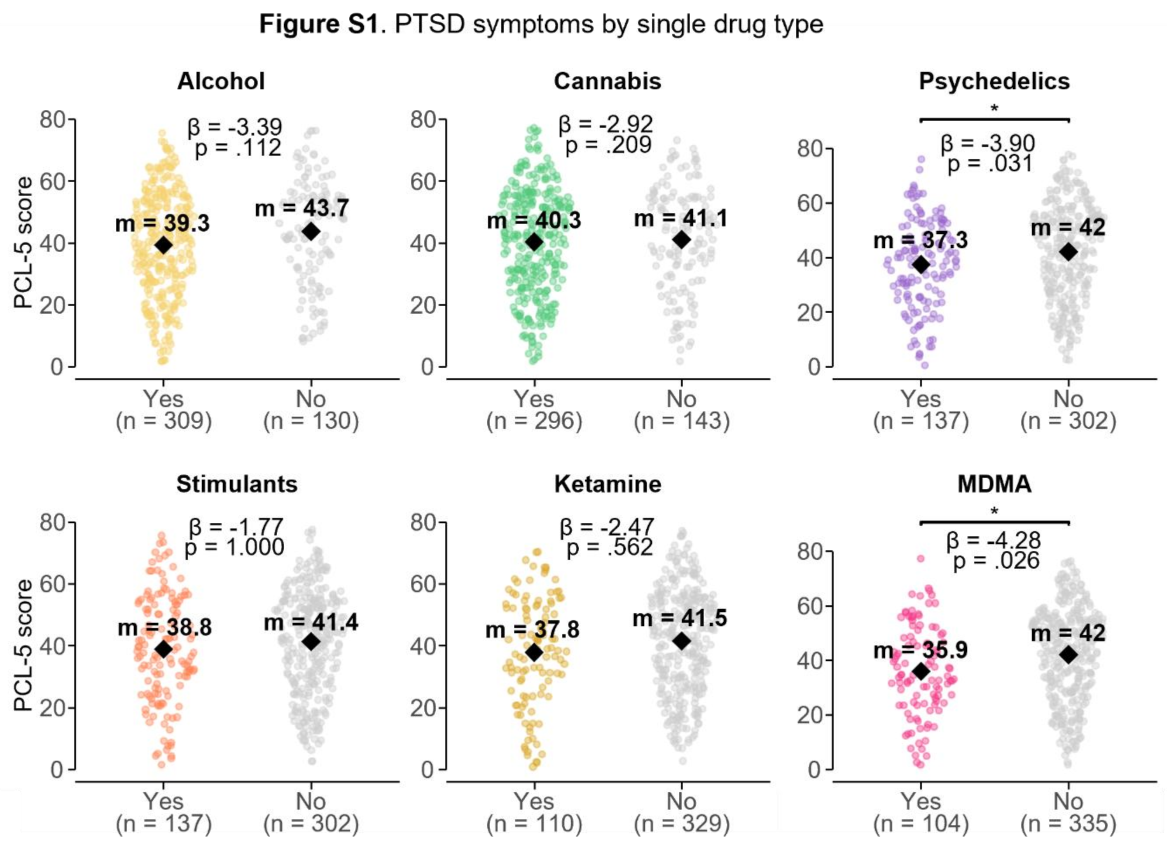


**Supplementary Figure S1.** PTSD symptoms by single drug type. PCL-5 at TP2 in users and non-users of each drug. For each drug, participants who used it in the month before TP2 (Yes; regardless of other drugs used) are compared with those who did not (No). Points are participants; diamonds and labels show observed means. **β** is the regression coefficient (Yes − No, PCL-5 points) adjusted for sex, age, months since October 7 and TP1 PCL-5; p-value is Bonferroni-corrected across the six drugs (* p<.05).

**Figure S2.** PTSD symptoms by single drug type compared to no use survivors

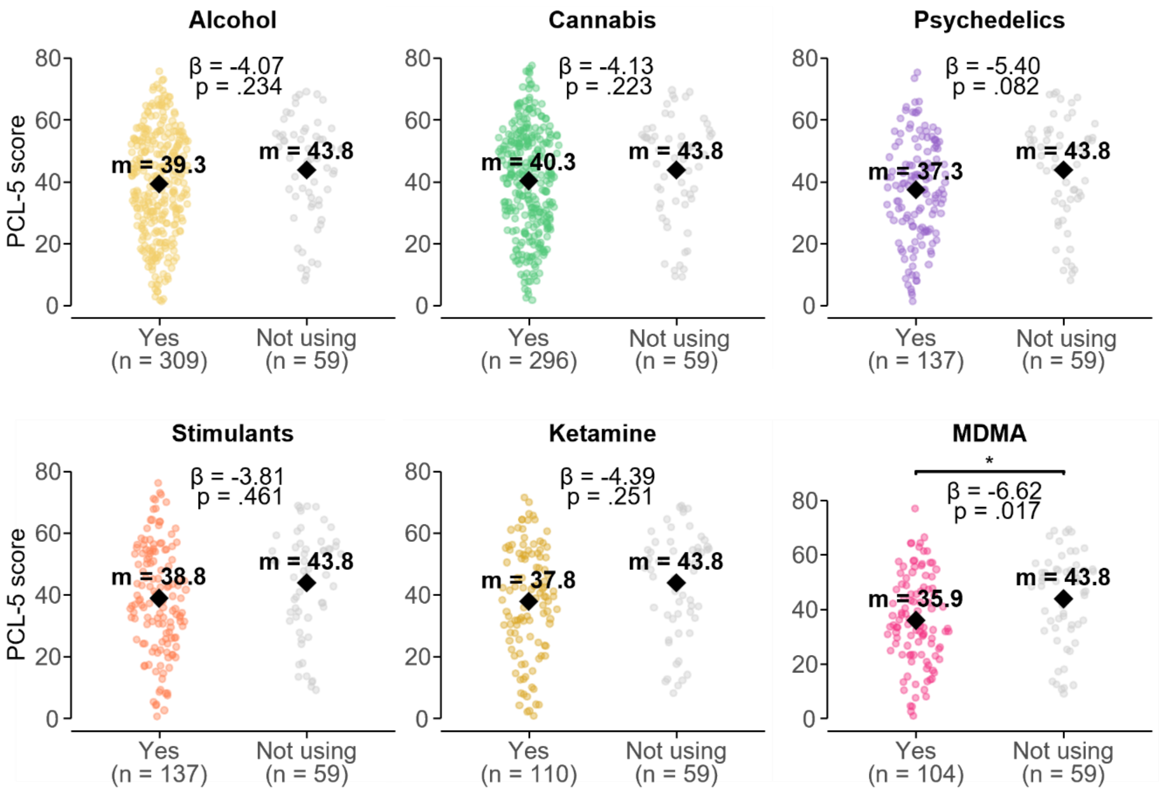


**Supplementary Figure S2.** PTSD symptoms by single drug type compared to no use survivors. Not using = not using any drug. PCL-5 at TP2 in users of each drug compared with abstinent participants. Users of each drug (Yes; regardless of other drugs used) are compared with participants reporting abstinence from all drugs (n=59). Points, means, **β** and p as in Supplementary Figure S1.

**Figure S3.** Frequency of use per month for low and high drug load class (Bars show % of users)

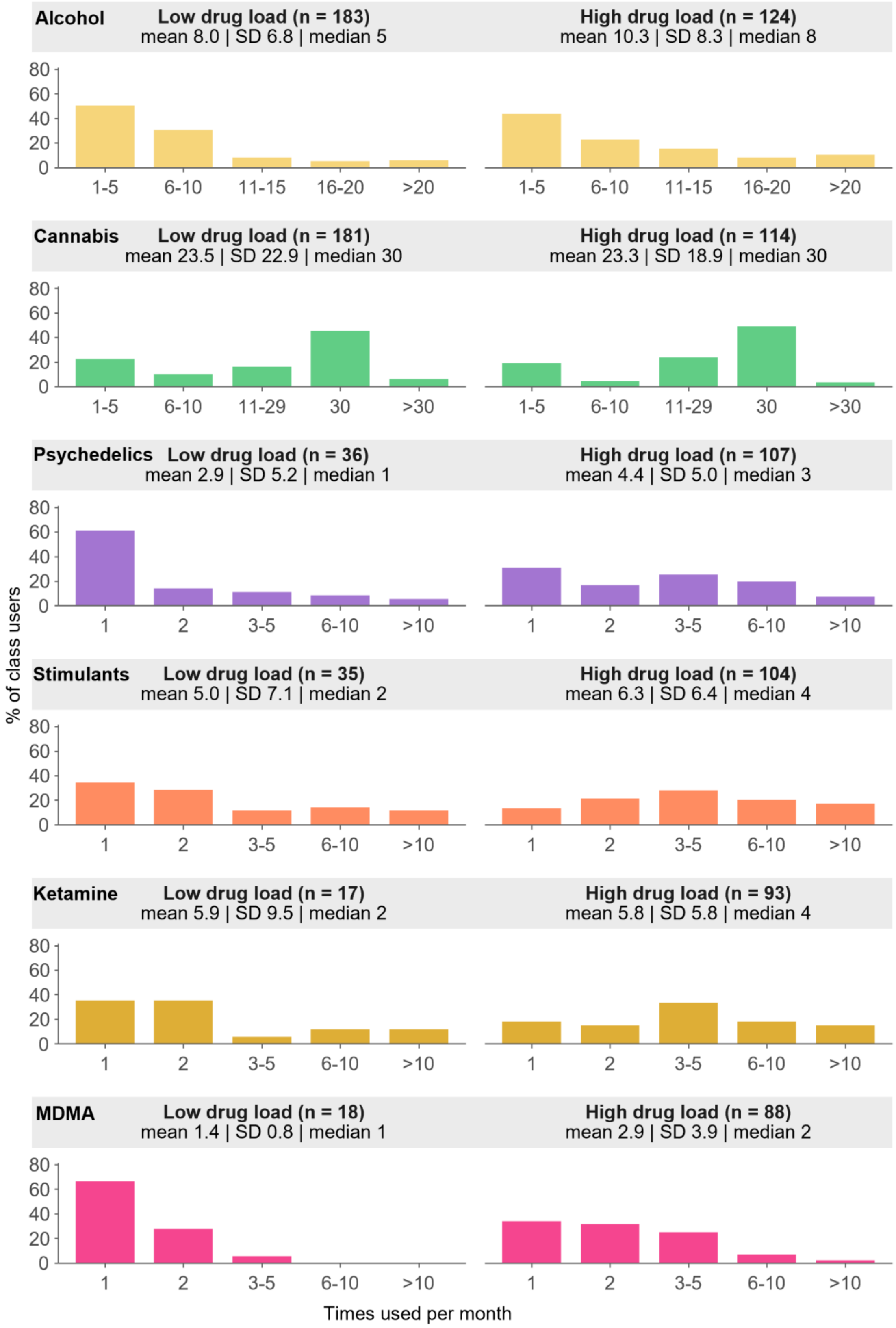

**Supplementary Figure S3.** Frequency of use by drug-load class. For each drug, the percentage of the class's users reporting each frequency (times used per month), shown separately for the Low drug load and High drug load classes (the Low-load and High-load classes in the text; users only). Panel headers give the number of users and the mean, SD and median times used per month in each class. Percentages are relative to the class's users of the drug, which accounts for the different class sizes.

**Figure S4.** Frequency of use per month for recreational and self-medication classification (Bars show % of users)

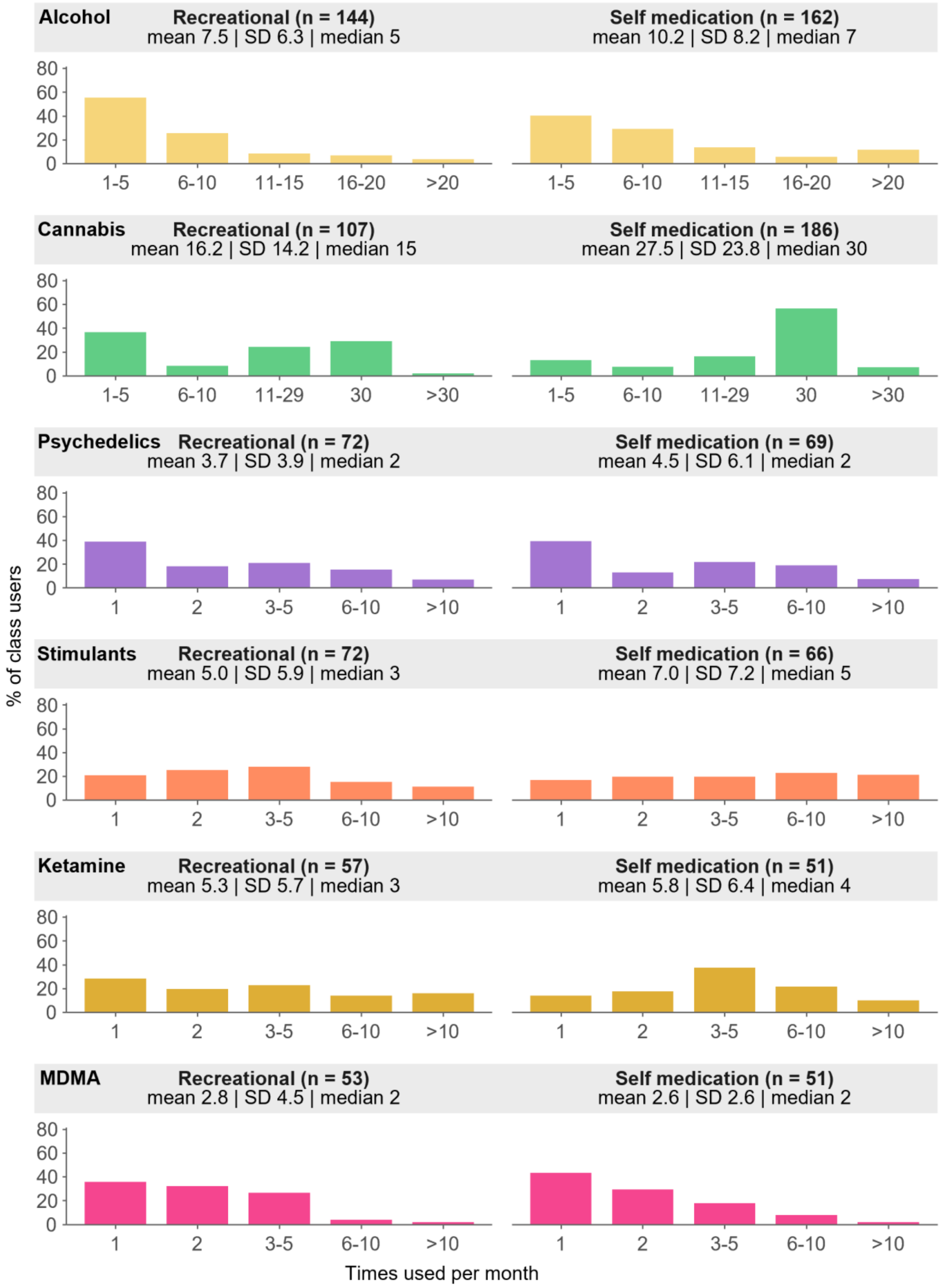

**Supplementary Figure S4.** Frequency of use by motive class. As in Supplementary Figure S3, for the Recreational and Self-medication classes.

**Figure S5.** Frequency of use by combined class (users only)
(Bars show % of users)

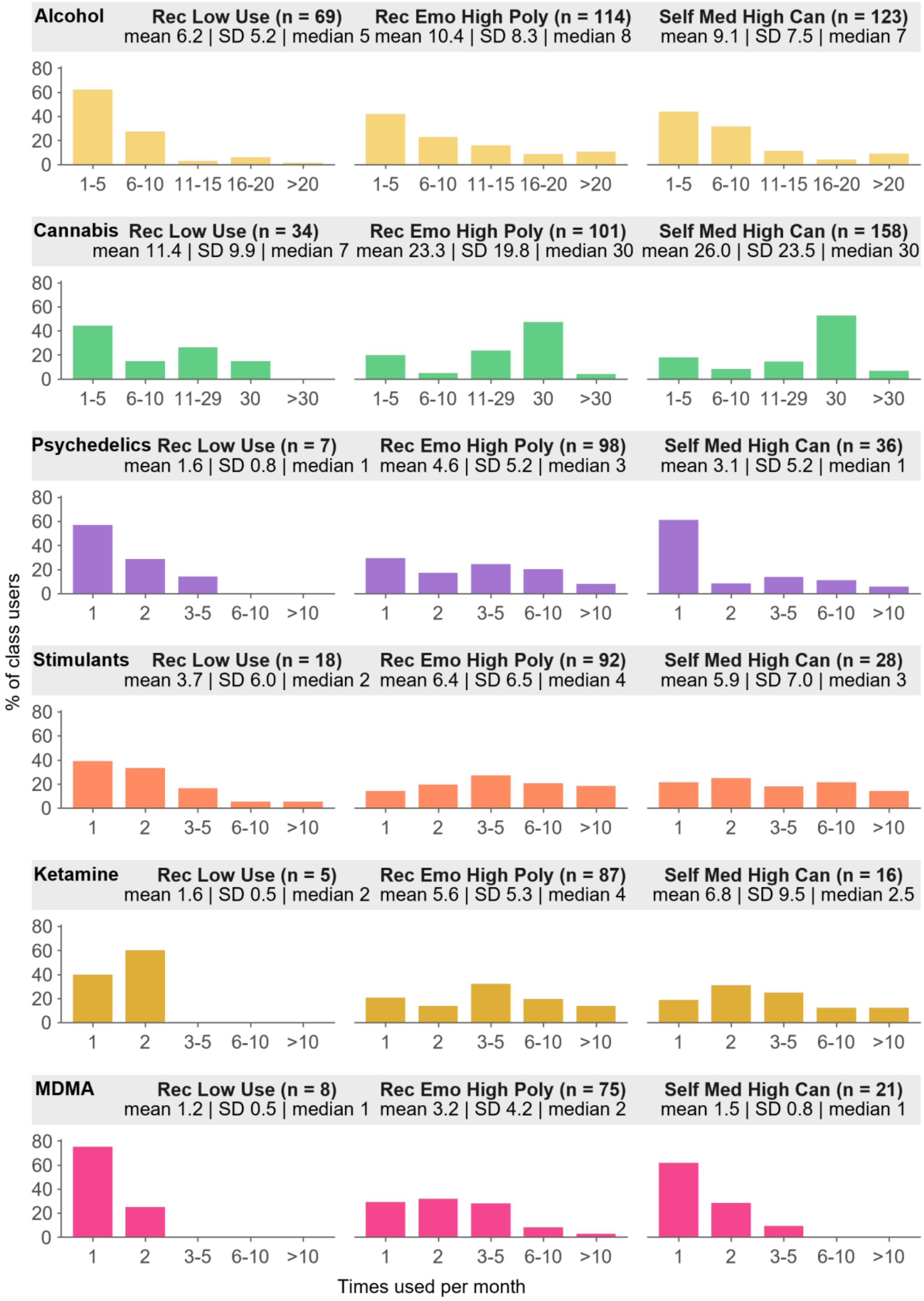

**Supplementary Figure S5. Frequency of use by combined class.** As in Supplementary Figure S3, for the Rec Low Use, Rec Emo High Poly and Self Med High Can classes.

**Figure S6**. PTSD symptoms by latent class adjusted for TP1 PCL-5

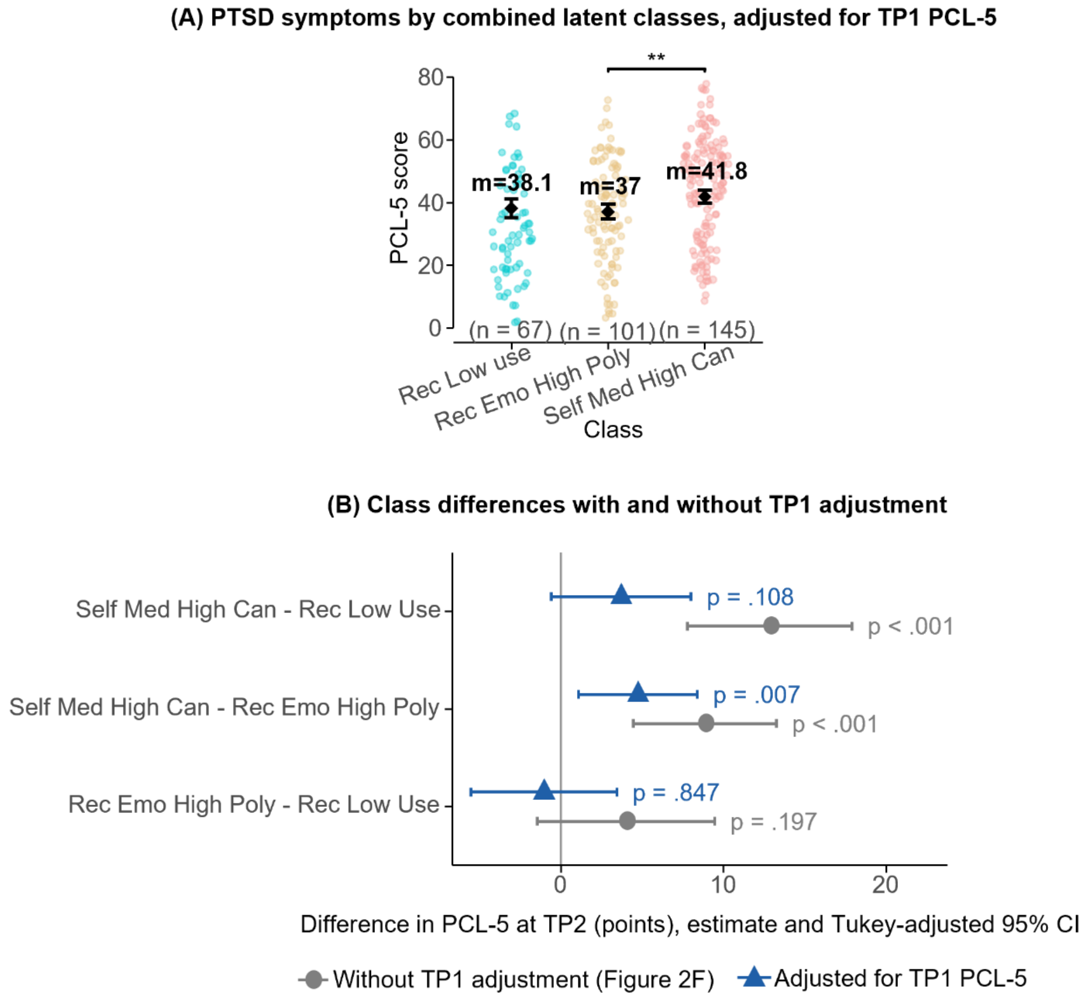


**Supplementary Figure S6.** Effect of adjusting for baseline (TP1) PTSD symptoms on the combined-class comparison. (A) PCL-5 at TP2 by combined latent class (n=313: Rec Low Use, 67; Rec Emo High Poly, 101; Self Med High Can, 145). Points are participants; diamonds and labels show estimated means, and error bars their 95% CI, adjusted for TP1 PCL-5, sex, age and time since the event; the bracket shows the Tukey-adjusted pairwise contrast (** p<.01; Table S1). (B) Pairwise class differences in PCL-5 at TP2 (estimate and Tukey-adjusted 95% CI) without adjustment for TP1 PCL-5 (grey circles; the model plotted in Figure 2F) and with adjustment (blue triangles). Differences are first-named minus second-named class; p-values are Tukey-adjusted.

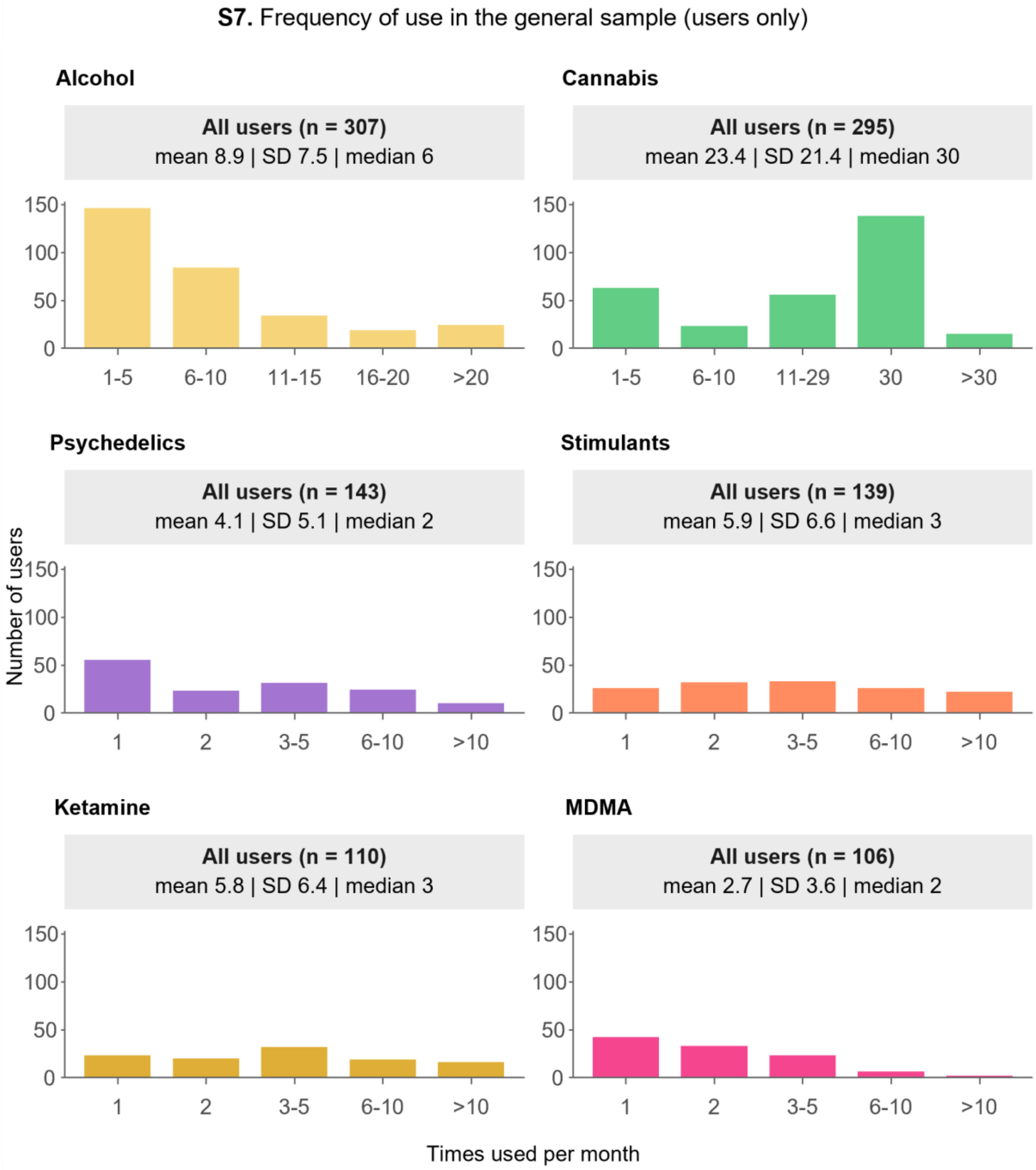


**Supplementary Figure S7.** Frequency of use in the general sample (users only). Distribution of the number of times each drug was used per month among users who reported a numeric frequency (alcohol, n=307; cannabis, n=295; psychedelics, n=143; stimulants, n=139; ketamine, n=110; MDMA, n=106). Bars show the number of users at each reported frequency; panel headers give the mean, SD and median of times used per month.
